# No SARS-CoV-2 in expressed prostatic secretion of patients with coronavirus disease 2019: a descriptive multicentre study in China

**DOI:** 10.1101/2020.03.26.20044198

**Authors:** Weihe Quan, Jun Chen, Zhigang Liu, Jinfei Tian, Xiangqiu Chen, Tao Wu, Ziliang Ji, Jinqi Tang, Hao Chu, Haijia Xu, Yong Zhao, Qingyou Zheng

## Abstract

**Purpose:** The aim of the present study was to assess whether SARS-CoV-2 can be detected in the expressed prostatic secretion (EPS) of patients with corona virus disease 2019 (COVID-19).

**Methods:** 18 cases of COVID-19, and 5 suspected cases, were selected from three medical centers to detect the RNA expression of SARS-CoV-2 in their EPS with RT-PCR.

**Results:** Results were negative in all EPS samples for SARS-CoV-2 of suspected and confirmed patients.

**Conclusions:** No SARS-CoV-2 was expressed in EPS of patients with COVID-19.

## Introduction

Since the first outbreak of COVID-19 in Wuhan city, December 2019, about 78,000 people have been infected in China. The causative agent of this disease, now called SARS-CoV-2, can cause acute respiratory distress syndrome, and the risk of death is relatively high^1^. The patients who were asymptomatic infected by SARS-CoV-2, constitute the main source of infection. The main routes of transmission are respiratory droplets and close contact^2^. In addition to secreta of respiratory tract, the researchers had isolated successfully the virus in the urine^3^, feces^4^ and conjunctiva^5^ of patients. It suggested that the virus may had multiple transmission routes. Based on the above studies, we propose the hypothesis that prostate may be a target organ of SARS-CoV-2, and that the RNA of virus expresses in the EPS of confirmed and suspected patients alike.

## Methods

### Study design and patients

From February 25 to March 13, 2020, 18 confirmed cases and 5 suspected cases of COVID-19 were selected from three medical centers to detect the RNA of virus expression in their EPS (Table 1). The confirmed cases in this study were SARS-CoV-2 positive in throat swabs; the suspected cases, while showing symptoms like fever and cough, were SARS-CoV-2 negative in the throat swabs twice with an interval of at least 24 hours. Cases of COVID-19 were diagnosed based on the Diagnosis and Treatment of COVID-19 (trial version 7) published by the National Health Commission of China. The three medical centers, Shenzhen Hospital of Southern Medical University, Wuhan Third Hospital (Hospital-Tongren) of Wuhan University and Xiangyang Central Hospital, are all specially designated by the Chinese government to treat COVID-19. The prevalence ranks from low to high in these three areas.

**Table 1.** Patient characteristics. BPH: benign prostatic hyperplasia; CP: chronic prostatitis; CRF: chronic renal failure; DM: diabetes mellitus; TCM: traditional Chinese medicine.

|  | **Confirmed patients** | **Suspected patients** |
| --- | --- | --- |
|  | n=18 | n=5 |
| **Throat swab (+)** | 100% | 0% |
| **Age,year** |  |  |
| Mean(SD) | 60.3 ± 15.3 | 45.6 ± 14.7 |
| Range |  |  |
| 20-39 | n=3 | n=3 |
| 40-59 | n=3 | n=1 |
| >60 | n=12 | n=1 |
| **Days of illness** |  |  |
| <1 week | n=8 | n=3 |
| 1-2 weeks | n=5 | n=1 |
| >2 weeks | n=5 | n=1 |
| **Urinary symptoms** |  |  |
| Frequency | n=2 | n=0 |
| Urgency | n=2 | n=0 |
| Pain | n=2 | n=0 |
| Dysuria | n=3 | n=0 |
| Hematuria | n=3 | n=0 |
| **Urinalysis** |  |  |
| WBC + | n=6 | n=0 |
| RBC + | n=5 | n=3 |
| Proteinuria | n=4 | n=2 |
| **Complication** |  |  |
| BPH | n=5 | n=1 |
| CP | n=4 | n=1 |
| Urolithiasis | n=9 | n=3 |
| CRF | n=2 | n=2 |
| Hypertension | n=8 | n=1 |
| DM | n=6 | n=1 |
| **Treatment** |  |  |
| Oxygen | n=12 | n=3 |
| Antibiotic | n=15 | n=3 |
| Antiviral | n=16 | n=4 |
| TCM | n=16 | n=3 |

The study was approved by the ethics committee of each hospital (NYSZYYEC20200008, KY2020-022, 2020-005), and has been registered in Chinese Clinical Trial Registry (ChiCTR2000030756). The patient consent was obtained.

### Data collection

All the samples were collected by trained medical professionals massaging the prostate through anus according to the published guidelines^6^ at the three centers. The EPS swab was saved in the virus sampling kit (Bangshuo, Guangzhou, China) immediately, All the samples were tested for SARS-CoV-2 with the recommended Kit (BGI, Shenzhen, China) from Chinese Center for Disease Control and Prevention (CDC), following WHO guidelines for real-time PCR. RT-PCR testing was performed according to the recommended protocol.

### Statistical analysis

SPSS software (version 20.0) was used for statistical analysis.

## Results

All 23 EPS samples of suspected and confirmed patients tested negative for the RNA of SARS-CoV-2 regardless by various grouping methods, such as days of illness, infected symptoms of urinary system (frequency, urgency and pain of urination), complication (chronic prostatitis, benign prostatic hyperplasia), or treatment method (antibiotic, antiviral, traditional Chinese medicine). Two cases among 18 confirmed patients. their EPS samples was negative on the day of discharge when their throat swabs turned negative and the clinical symptoms disappeared.

## DISCUSSION

The pathogen that causes the COVID-19 has been identified as a novel enveloped RNA beta corona virus 2^7^. As a phylogenetic similar to SARS-CoV, it has currently been named severe acute respiratory syndrome corona virus 2 (SARS-CoV-2). Human beings has no immunity to the virus and is susceptible. The incubation period lasts usually 1 to 14 days. Fever and cough are the dominant symptoms,mostly occur at 3 to 7 days after infection. Gastrointestinal and urological symptoms are rare. A few patients are accompanied by symptoms such as nasal obstruction, runny nose, sore throat, myalgia and diarrhea. Severe patients often have dyspnea and hypoxemia one week after the onset of the disease, then rapidly progress to acute respiratory distress syndrome, septic shock, metabolic acidosis, coagulation dysfunction and multiple organ dysfunction syndrome^2^.

Conventional routes of transmission of SARS-CoV-2 consist of respiratory droplets and direct contact. It is possible to propagate through aerosols when exposed to high concentration aerosols for a long time in a relatively closed environment^4^. From the China-WHO investigation on COVID-19, it reported that SARS-CoV-2 was an animal virus, and that air transmission was not considered as the main mode. it indicated that the interpersonal transmission mainly occurred within households in China. So far SARS-CoV-2 has been detected in the gastrointestinal tract, saliva, and urine, so these routes of potential transmission need to be investigated.

It has been reported that SARS-CoV-2 shares 76% amino acid sequence identity with severe acute respiratory syndrome corona virus (SARS-CoV) and is likely to use the same receptor, angiotensin-converting enzyme 2 (ACE2), for entry into target host cells^8,9^, The binding affinity of SARS-CoV-2 with its receptor is more 10 to 20 times higher than ones of else corona virus, like SARSr-CoV and MERSr-CoV^10^. Investigation of the expression pattern of ACE2 in adult human testis at the level of single-cell transcriptomes indicates that ACE2 is predominantly enriched in spermatogonia, leydig and sertoli cells. Further research shows that enrichment of ACE2 in leydig and sertoli cells has a significant higher frequency compared with ACE2-expressing cells in type II alveolar epithelial cells (AT2) in human lung (4.25% vs 1.40%)^8^. These evidences suggested that the human testis was a high-risk organ vulnerable to SARS-CoV-2 infection that may result in spermatogenic failure^11^.

ACE2 receptor is also expressed in human kidney. Recently, Academician Zhong Nanshan’s team isolated SARS-CoV-2 from urine samples of a patient with COVID-19. A latest study has showed that ACE2 was not only highly expressed in lung AT2 cells but also in absorptive enterocytes from ileum and colon, has indicated that digestive system was a potential route for SARS-CoV-2 infection^12^. It may be a reason by fecal-oral transmission. It has reported that the result of anus swab of SARS-CoV-2 turned positive again in a few of discharged patients during the follow-up^13^. These patients need to be closely monitored for infectivity.

All above evidences show that the respiratory system, digestive system and urogenital system are all the target organs of SARS-CoV-2^14,15^. It is inferred that the testis and prostate of patients with COVID-19 are also vulnerable to SARS-CoV-2, so we collect the EPS of confirmed and suspected patients to test. According to our results, there is no any affirmative evidence of the virus expressed in EPS samples, which might indicate that prostate may be not a target organ of SARS-CoV-2, and the virus will not appear in the EPS regardless of the throat swab results. However, some limitations should be noted. First, although those samples were collected from three medical centers with different prevalence rates, EPS were tested only in mild and common patients, excluding samples of severe patients. Second, semen samples were not obtained, due to some restrictions like non-private ward environment of mobile cabin hospitals, anxiety during isolation period, so the safety of sexual intercourse still remains to be known. Third, our sample size is relatively small, with only 60 cases. In the next study, large samples and long-term follow-up are needed to assess whether sexual intercourse is a potential route for virus transmission.

## Data Availability

With the permission of the corresponding authors, we can provide participant data without names and identifiers, but not the study protocol, statistical analysis plan, or informed consent form. Data can be provided after the article is published. Once the data can be made public, the research team will provide an email address for communication. The corresponding authors have the right to decide whether to share the data or not based on the research objectives and plan provided. 

## Contributors

WQ, QZ, and J Tian made substantial contributions to the study concept and design. WQ, JC, and ZL took responsibility for obtaining written consent from patients, obtaining ethical approval, and confirming data accuracy. XC, TW, HC, HX and ZJ took responsibility for collecting samples and data. ZY tested some samples. J Tian, XC and TW made substantial contributions to data acquisition, analysis, and interpretation. J Tian, JC, WQ and ZL participated in drafting the manuscript, and revising it on the basis of reviewers ‘ comments. WQ, J Tian and QZ revised the final manuscript. WQ, JC, ZL and J Tian contributed equallyand share co-first authorship.

## Reference

1. Chen, N., Zhou, M., Dong, X., Qu, J., Gong, F., Han, Y., Qiu, Y., Wang, J., Liu, Y., Wei, Y. et al. (2020) Epidemiological and clinical characteristics of 99 cases of 2019 novel coronavirus pneumonia in Wuhan, China: a descriptive study. Lancet

2. Diagnosis and treatment of COVID-19 (trial version 7), National Health Commission of the People’s Republic of China. http://www.nhc.gov.cn/yzygj/s7653p/202003/46c9294a7dfe4cef80dc7f5912eb1989.shtml

3. Guan W, Ni Z, Hu Y, et al. Clinical characteristics of coronavirus disease 2019 in China. N Engl J Med. DOI:10.1056/NEJMoa2002032

4. Wei-jie Guan, Ph.D., Zheng-yi Ni, M.D., Yu Hu, M.D., Wen-hua Liang, Ph.D., Chun-quan Ou, Ph.D., Jian-xing He, M.D., Lei Liu, M.D., Hong Shan, M.D., Chun-liang Lei, M.D., David S.C. Hui, M.D., Bin Du, M.D., Lan-juan Li, M.D., Guang Zeng, M.Sc., Kwok-Yung Yuen, et al. Clinical characteristics of 2019 novel coronavirus infection in China[EB/OL]. https://www.medrxiv.org/content/10.1101/2020.02.06.20020974v1.

5. Xia J, Tong J, Liu M, Shen Y, Guo D. Evaluation of coronavirus in tears and conjunctival secretions of patients with SARS-CoV-2 infection. J Med Virol.2020 Feb 26. doi:10.1002/jmv25725. [Epub ahead of print]

6. Centers for Disease Control and Prevention. Interim guidelines for collecting,handling,and testing clinical specimens from patients under investigation (PUIs) for 2019 novel coronavirus (2019 ? nCoV). https://www.cdc.gov/coronavirus/2019-nCoV/guidelines-clinical-specimens.html.

7. Lu R, Zhao X, Li J, et al. Genomic characterisation and epidemiology of 2019 novel corona virus: Implications for virus origins and receptor binding[J]. Lancet, 2020

8. Yu Zhao, Zixian Zhao, Yujia Wang, Yueqing Zhou, Yu Ma and Zuo, W. (2020) Singlecell RNA expression profiling of ACE2, the putative receptor of Wuhan 2019-nCov. bioRxiv

9. Zhou P, Yang X L, Wang X G, et al. A pneumonia outbreak associated with a new coronavirus of probable bat origin[J]. nature, published on February o3,2020. DOI: 10.1038/s41586-020-2012-7

10. Xu X, Chen P, Wang J, et al. Evolution of the novel coronavirus from the ongoing Wuhan outbreak and modeling of its spike protein for risk of human transmission[J]. SCIENCE CHINA-LIFE SCIENCES, 2020.

11. Zhengpin Wang,Xiaojiang Xu. scRNA-seq profiling of human testes reveals the presence of ACE2 receptor, a target for SARS-CoV-2 infection, in spermatogonia, Leydig and Sertoli cells.doi: 10.20944/preprints202002.0299.v1

12. Zhang H, Kang Z, Gong H, et al. The digestive system is a potential route of 2019-nCoV infection: A bioinformatics analysis based on single-cell transcriptomes[Z]. Biorxiv, 2020, https://doi.org/10.1101/2020.01.30.927806

13. Zou jingbo,Zhou yang,Qiao jie, et al. A case report of virus nucleic acid positive in feces of patients with covid-19 after treatment in Chongqing[J/OL]. Chinese Journal of Virology. https://doi.org/10.13242/j.cnki.bingduxuebao.003653.

14. Hao Zhang, Zijian Kang, Haiyi Gong, D. Xu, Jing Wang, Zifu Li, Xingang Cui, Jianru Xiao, Tong Meng, Wang Zhou et al. (2020) The digestive system is a potential route of 2019-nCov infection: a bioinformatics analysis based on single-cell transcriptomes. bioRxiv.

15. Xin Zou, Ke Chen, Jiawei Zou, Peiyi Han, Jie Hao and Han, Z. (2020) The single cell RNA seq data analysis on the receptor ACE2 expression reveals the potential risk of different human organs vulnerable to Wuhan 2019 nCoV infection. Frontiers of Medicine.

